# Prenatal maternal distress during the COVID-19 pandemic and its effects on the infant brain

**DOI:** 10.1101/2021.10.04.21264536

**Authors:** Kathryn Y. Manning, Xiangyu Long, Dana Watts, Lianne Tomfohr-Madsen, Gerald F. Giesbrecht, Catherine Lebel

**Affiliations:** Department of Radiology, University of Calgary; Alberta Children’s Hospital Research Institute; Hotchkiss Brain Institute, University of Calgary; Department of Psychology, University of Calgary; Department of Pediatrics, University of Calgary; Department of Educational and Counselling Psychology, University of Calgary; Department of Community Health Sciences, University of Calgary

## Abstract

The COVID-19 pandemic has caused elevated distress in pregnant individuals, which has the potential to impact the developing infant. In this study, we examined anxiety and depression symptoms during the pandemic in a large sample of pregnant individuals (n=8602). For a sub-sample of participants, their infants underwent magnetic resonance imaging (MRI) at 3-months of age to examine whether this prenatal maternal distress was associated with infant brain changes. We found significantly elevated prenatal maternal distress compared to pre-pandemic rates, with 47% and 33% of participants reporting clinically significant symptoms of anxiety and depression, respectively. Importantly, we identified social support as a protective factor for clinically elevated prenatal maternal distress. We found significant relationships between prenatal maternal distress and infant amygdala-prefrontal microstructural and functional connectivity and demonstrate for the first time that social support moderates this relationship. Our findings suggest a potentially long-lasting impact of the COVID-19 pandemic on children and show that social support acts as a protective factor not just for pregnant individuals, but also for their developing infants. These findings provide timely evidence to inform clinical practice and policy surrounding the care of pregnant individuals and highlight the importance of social support.

## Introduction

Prenatal maternal distress, defined as elevated symptoms of anxiety and/or depression, can have immediate negative effects on pregnant individuals as well as the rapidly developing and vulnerable fetus. Prenatal maternal distress is associated with preterm birth as well as long-term risks for behavioural and mental health problems in children^1–3^. Alterations to the developing infant brain, especially within the limbic system, likely underlie compromised behavioural development^4–7^. In particular, infants exposed to higher prenatal stress have larger amygdala volumes, disrupted white matter connectivity, and functional connectivity changes between the amygdala and prefrontal regions^8–10^. Further, amygdala-prefrontal pathways mediate the relationship between maternal prenatal depression and externalizing behaviours (hyperactivity, aggression) in children^11^, showing that brain structure is a mechanism via which prenatal maternal distress can impact children’s behavioural development.

The COVID-19 pandemic has had a profound and prolonged effect on the mental health of pregnant individuals. Pregnant individuals have faced fear for themselves and their developing babies, disruptions to prenatal care (including lack of partner support)^12^, reduced access to services, and loss of support from friends and family. Rates of psychological distress in pregnant individuals have more than tripled during the pandemic^13–15^, compared to rates pre-pandemic^16,17^. Previous studies of children born during natural disasters demonstrate the long-term consequences of prenatal distress on brain development^18,19^ and behaviour^20^, raising significant concerns about how the generation of children born during the COVID-19 pandemic will be affected. Data is emerging that demonstrates deficits in early infant and child cognitive development^21^.

Social support can moderate the effects of prenatal distress on maternal HPA axis activity^22^ and has been shown to reduce psychological distress during pregnancy^16,23,24^. However, it is unclear whether social support can disrupt the transmission of prenatal maternal distress to infant brain development. Understanding how prenatal maternal distress affects brain development, including potential modifiable risk and protective factors such as social support, will aid in identifying children most at risk and inform policy recommendations to rapidly benefit families.

Here, we measured prenatal depression, anxiety, and social support in a very large sample of pregnant individuals across Canada during the COVID-19 pandemic. We tested associations between prenatal maternal distress and social support. In a subset of infants, we examined how prenatal distress was related to infant brain structure and function, and whether social support moderated the associations.

## Methods

### Participants

The Pregnancy during the COVID-19 Pandemic (PdP) study^25^ is a Canada-wide study that recruited pregnant individuals between April 2020-April 2021 to assess life changes, physical and mental health during the pandemic. The University of Calgary Conjoint Health Research Ethics Board (CHREB) approved this study and participants provided written consent. Pregnant individuals were eligible if they were living in Canada, ≥17 years of age, <35 weeks’ gestation at intake, and able to read and write in English or French. Full details about the study are provided elsewhere^25^. Here, we use data from the first intake survey. 8602 participants (mean age=32.0 +/- 4.4 years, range 17-49 years; mean gestation=20.7+/- 8.6, range 3-35 weeks) completed the mental health and social support measures and were included in this study. We also gathered information on maternal age, education, and ethnicity using self-selection tools through the online survey (see eTable 1).

### Mental Health and Social Support

The Edinburgh Depression Scale (EPDS)^26^ was used to assess depression symptoms and the Patient Reported Outcomes Measurement Information System (PROMIS) Anxiety^27^ scale to assess anxiety symptoms, and the Social Support Effectiveness Questionnaire (SSEQ)^28^ to assess the amount and quality of social support provided by a partner or close friend/family member. Clinically elevated depression symptoms were defined as a score of >12 on the EPDS and clinically elevated anxiety was defined as scores > 59.

### Image Acquisition

PdP participants who delivered full term infants (≥37 weeks) in the Calgary area were invited to participate in neuroimaging when infants were 3 months of age. Participants received a stipend for participating in the imaging portion of the study. Children with major birth complications (e.g., hypoxic-ischemic encephalopathy), diagnosed genetic or neurological conditions associated with significant cognitive impairment, congenital anomalies, or contraindications to MRI were excluded. 75 infants (92 +/- 14 days old) underwent brain imaging on a GE 3T MR750w using a 32-channel head coil at the Alberta Children’s Hospital. Infants were scanned during natural sleep while placed in an inflatable MedVac infant scanning bed. The imaging protocol included T1-weighted imaging (TE = 2200 ms, TR = 5200 ms, TI = 540 ms, FOV = 1900 mm, matrix = 512 × 512, bandwidth = 41.67, voxels 0.37 × 0.37 × 1 mm^3^, flip angle = 12°, 136 slices, total time=X), diffusion tensor imaging (DTI) (30 directions at b = 700 s/mm^2^, 5 b=0 s/mm^2^ images, TR = 8500 ms, TE = 99.4 ms, FOV = 1600 mm, matrix = 256 × 256, voxel 0.625 × 0.625 × 2 mm^3^, 49, slices total time = 5:06) and resting state functional MRI (rs-fMRI) (TR = 2000 ms, TE = 30 ms, FOV = 1920 mm, matrix = 64 × 64, voxel 3.6 × 3.6 × 3.6 mm^3^, flip angle = 60°, 37 slices, total time = 8:10).

### Image Analysis

ExploreDTI^29^ was used to process the diffusion data, including signal drift, Gibbs ringing, eddy currents and head motion corrections. Participants who awoke early during the scanning protocol and were removed from the scanner before completing the diffusion portion of the protocol or those with excessive motion and compromised diffusion data determined with manual visual check were not included in data analyses. Whole brain whiter matter tractography was performed using semi-automated deterministic streamline tractography with fractional anisotropy (FA) > 0.1 and angle < 30 degrees. The bilateral uncinate fasciculus and fiber bundles connecting the amygdala to the prefrontal cortex were generated using our previous methods^11^. Average FA and mean diffusivity (MD) were calculated within each tract for each participant.

Rs-fMRI preprocessing was conducted using FSL^30^ and included brain extraction, motion correction, 5mm smoothing, and high pass temporal filtering. Participants were excluded if they awoke early during the scanning protocol and were removed from the scan before completing the functional portion of the protocol or if they had excessive motion (> 1mm relative mean displacement for > 120 volumes). Independent component analysis (ICA) denoising was used to regress non-neuronal components from the data and clean images were registered to an infant atlas template for neonates^31^. FEAT^32^ higher-level analysis was used to create an average amygdala connectivity map (Figure 2A) and identify regions of interest (superior orbitofrontal cortex and inferior frontal gyrus), delineated using the infant Automated Anatomical Labeling (AAL) atlas^31^. We tested intrahemispheric functional connectivity between these two prefrontal regions and the amygdala using a Pearson correlation in MATLAB.

### Statistical Analysis

Standard scores from the EPDS and PROMIS Anxiety measures were combined using a factor analysis in SPSS into a single factor to quantify prenatal maternal distress^5^. A general linear model was used to investigate the relationship between prenatal maternal distress and SSEQ, while controlling for maternal education and ethnicity. We also conducted a logistic regression analysis to determine if SSEQ score predicts clinically significant prenatal psychological distress.

For the infant imaging data, a general linear model was applied to examine the relationship between each MRI metric (i.e., rs-fMRI functional connectivity, FA, or MD) and prenatal maternal distress. We included maternal education, age of the infant at the time of the scan, infant sex, and SSEQ in our model as follows:

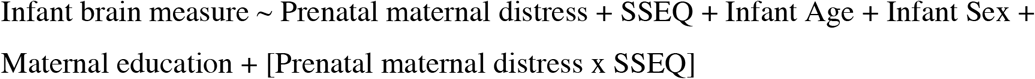

where the relationship between MRI measures and prenatal maternal distress were tested for significance at *p* < 0.05. Results were corrected for multiple comparisons using false-discovery rate (FDR) correction based on 12 general linear models (4 FA measures, 4 MD measures and 4 functional connectivity measures). The interaction term between SSEQ and prenatal maternal distress was included to examine whether social support moderates the relationship between prenatal maternal distress and the developing infant brain. If the interaction was significant, post-hoc tests between high and low social support groups (based on the median i.e. above and below SSEQ=60) were used to further understand the relationships. If the interaction term was not significant, the model was re-run without it included.

### Data Availability

The COVID-19 PdP Study data, including maternal mental health measures and infant imaging data is available upon request.

## Results

### Prenatal maternal distress and social support

The average EPDS score was 10.4 +/- 5.3 (range: 0-30), with 33.4% participants demonstrating clinically elevated depression symptoms (scores > 12). The average PROMIS T-score was 58.5 +/- 8.0 (range: 36.3-82.7), with 47.1% of participants demonstrating clinically elevated anxiety symptoms. The average SSEQ score was 50.3 +/- 5.7 (range: 26-71).

Prenatal maternal distress was substantially elevated compared to pre-pandemic rates and significantly related to SSEQ (R = -0.3, *p* < 0.001, 95% CI: [-0.33, -0.29]), while controlling for maternal education and ethnicity (Figure 1). The logistic regression was also significant (T = - 22.3, *p* < 0.001, β = -0.06, 95% CI: [-0.06, -0.05]) where higher SSEQ was identified as a protective factor for clinically significant prenatal maternal distress.

**Figure 1:**
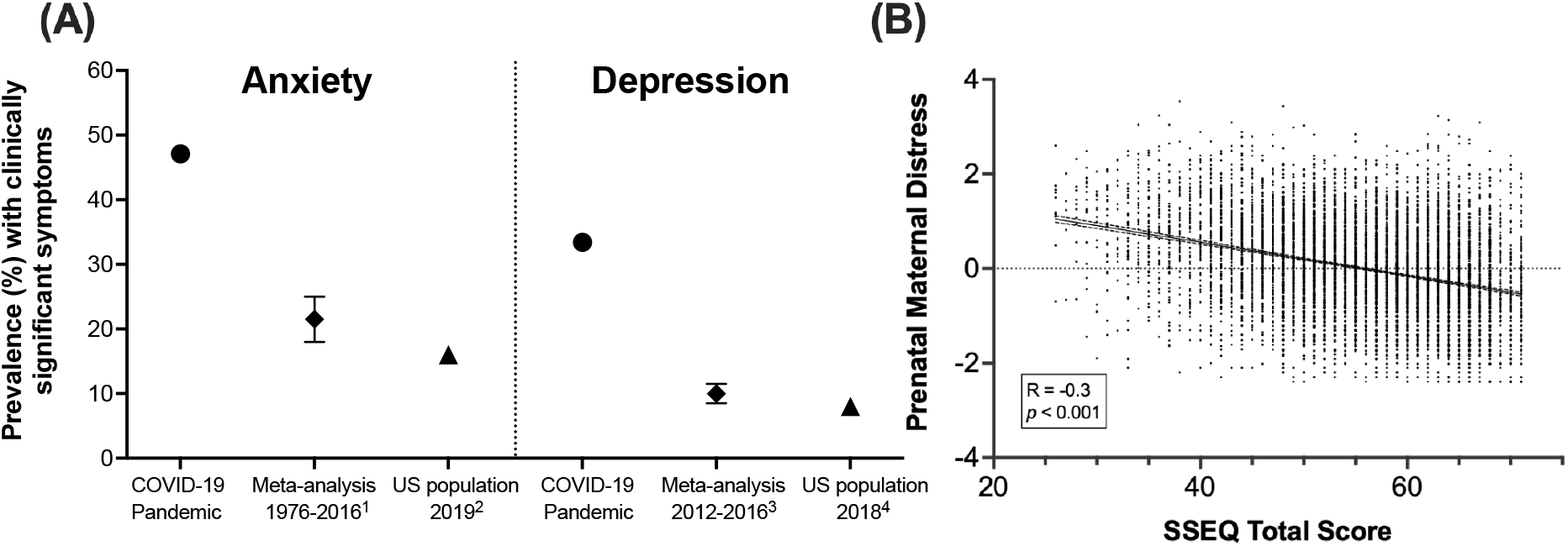
Elevated prenatal maternal distress and social support. (A) The percent of pregnant individuals from the current study (N = 8602) that had clinically significant symptoms of anxiety or depression compared to meta-analyses of pregnant individuals pre-pandemic (reference 1^33^ and reference 3^34^; 95% CI shown with error bars) and US general population norms (reference 2^35^ and reference 4^36^). (B) There was a negative relationship between SSEQ total score and prenatal maternal distress, while controlling for maternal education and ethnicity. Higher social support was associated with lower prenatal maternal distress.

**Figure 2:**
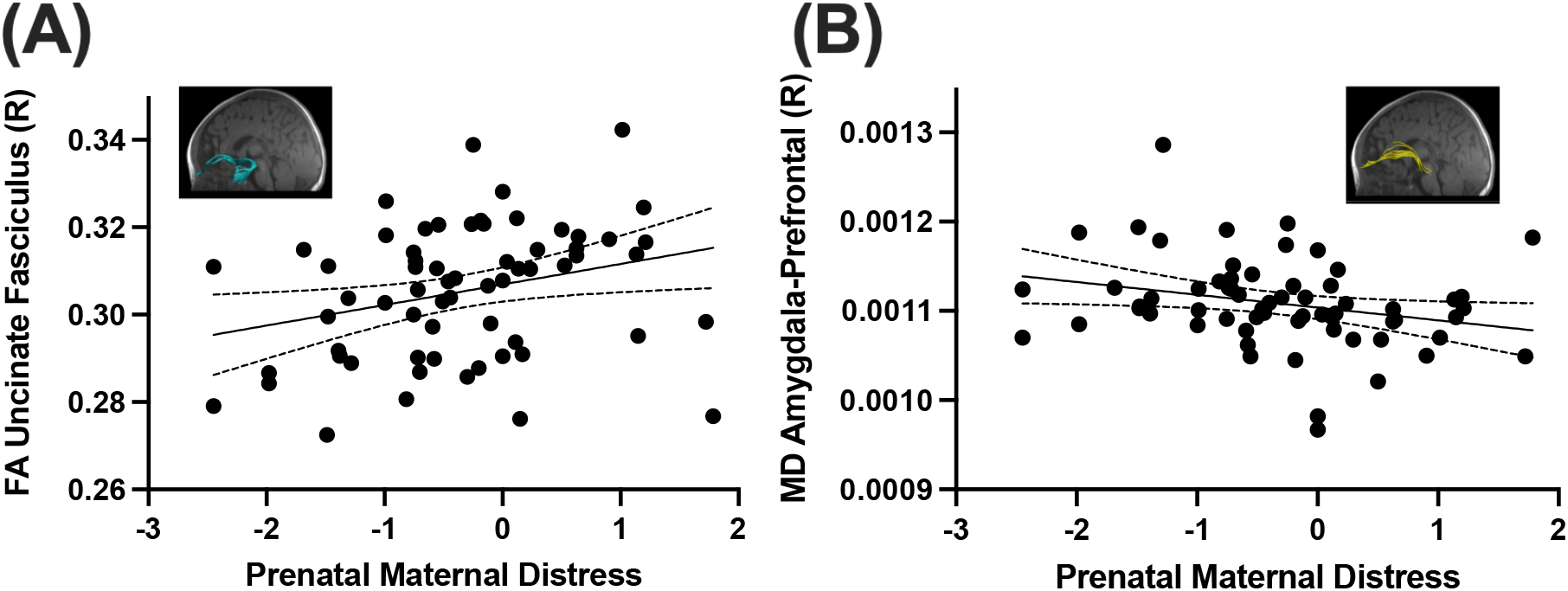
Prenatal maternal distress and infant brain microstructure. Prenatal maternal distress was significantly positively correlated with average fractional anisotropy (FA) in the right uncinate fasciculus (A), and negatively correlated with mean diffusivity (MD) in the right amygdala-prefrontal tract (B).

### Infant brain structure and function

A total of 75 infants participated in imaging at the Alberta Children’s Hospital between August 2020 and May 2021. Average EPDS and PROMIS Anxiety scores in this sub-sample were 8.6 +/- 5.3 and 54.7 +/- 12.4, respectively. These scores are significantly lower than the full survey sample (*p* < 0.01), but still elevated compared to pre-pandemic anxiety and depression levels. After image quality control (described in the methods section), we retained 63 DTI datasets (40M/23F, 92 +/- 14 days old). Of the 12 participants with incomplete/poor data, 6 participants awoke early and were unable to complete the protocol and 6 had excessive motion during the scan. After the functional imaging quality control, we retained 45 rs-fMRI datasets (30M/15F, 92 +/- 14 days old). Of the 30 participants with incomplete/poor data, 14 participants had excessive motion, 14 awoke before the rs-fMRI acquisition and did not complete the protocol, and 2 had a field of view error.

The SSEQ x prenatal maternal distress interaction terms were not significant for diffusion measures, so the model was run with this term removed (eTable 2). Average FA in the right uncinate fasciculus (T = 2.8, *p* = 0.008, β = 0.006, 95% CI: [0.002, 0.01]) and average MD in the right amygdala-prefrontal white matter tract (T = -2.6, *p* = 0.01, β = -0.0002, 95% CI: [-0.0003, - 0.00004]) were significantly related to prenatal maternal distress (Figure 2).

Functional connectivity between the right amygdala and right superior orbitofrontal cortex had a significant main effect of prenatal distress (T = -2.7, *p* = 0.01, β = -0.3, 95% CI: [-0.5, -0.08]) with a significant SSEQ x prenatal distress interaction (T = 2.3, *p* = 0.02, β = 0.004, 95% CI: [0.001, 0.008]) (Figure 3). Post-hoc tests revealed that pregnant individuals who reported lower quality social support (< 60) had a significant negative correlation between prenatal maternal distress and infant functional connectivity (R = -0.4, *p* = 0.04, 95% CI: [-0.7, -0.02]), and those who reported higher social support did not (R = -0.08, *p* = 0.7, 95% CI: [-0.08, 0.06]). In other words, higher prenatal maternal distress was associated with weaker amygdala-prefrontal functional connectivity when social support was low, but there was no association when social support was high.

**Figure 3:**
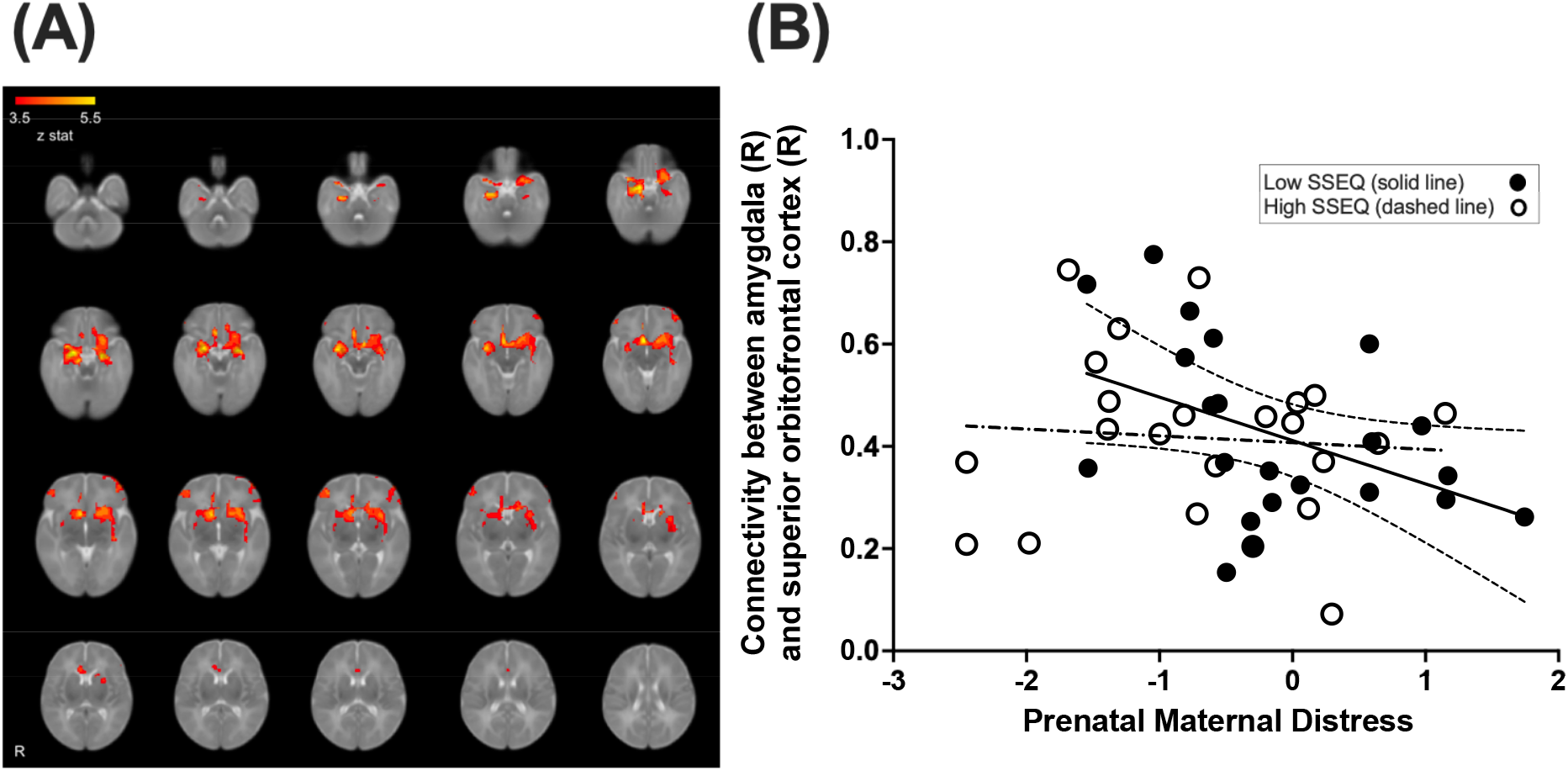
Prenatal maternal distress and infant brain functional connectivity. Average whole-brain functional connectivity of the amygdala is shown in infants (A). Functional connectivity from the right amygdala and the right superior orbitofrontal cortex demonstrated a significant interaction between SSEQ and prenatal maternal distress (B). The low SSEQ group demonstrated a significant negative correlation between maternal distress and connectivity (solid line) and the high SSEQ group did not (dashed line).

## Discussion

Here, in a very large cross-Canada sample of pregnant individuals, we show substantially elevated prenatal maternal distress that is associated with structural and functional brain alterations in their infants. One third and nearly half of pregnant individuals demonstrated clinically significant depression and anxiety symptoms, respectively. This is consistent with other studies around the world and demonstrates how substantially affected pregnant people have been during this pandemic^17^. The prevalence of clinically elevated depression and anxiety symptoms among this cohort has increased substantially in comparison to pre-pandemic distress levels among pregnant individuals^37^ and US population norms^27,36^ (see Figure 1). Importantly, prenatal psychological distress was moderated by social support where individuals with better quality and/or quantity of perceived social support reported lower symptoms of anxiety and depression. In a subsample of participants, we demonstrated that prenatal distress was associated with white matter microstructure and functional connectivity in the infant brain. Furthermore, we demonstrate for the first time that the relationship between prenatal distress and functional brain connectivity is moderated by social support, suggesting that social support in pregnancy may not only support mothers, but can also mitigate the intergenerational transmission of prenatal stress.

In the largest sample to date of pregnant individuals during the COVID-19 pandemic, our findings affirm previous research showing that a high level of perceived social support from a partner or another supportive person may buffer the severity of psychological symptoms among pregnant individuals^38,39^. Our results reflect the importance of strong social support for pregnant individuals, and the role of partners and others in maintaining healthy prenatal mental health, where we identified that social support was a protective factor for clinically elevated prenatal maternal distress. Partner support may have been especially important during the COVID-19 pandemic, when individuals were isolated from family and friends, as well as from the general community, due to government restrictions and fears of exposure to the virus. Partners may have had limited access to attend appointments, and some prenatal appointments were virtual instead of in-person. Changes in a support person attending the birth specifically have been related to elevated symptoms of prenatal anxiety and depression^12^.

In infants born during the pandemic, we showed both structural and functional alterations in amygdala-prefrontal connectivity associated with prenatal distress. Infants exposed to higher prenatal maternal anxiety and depression had higher FA and lower MD. Because FA generally increases and MD generally decreases across childhood^40^, this suggests premature development of the right amygdala-prefrontal white matter connections involved in emotion regulation.

Previous studies of infant brain microstructure and maternal prenatal depression have reported mixed findings, with some showing higher prenatal depressive symptoms related to lower neurite density^5^ and lower FA^9^ in 1-month and two-week old infants, respectively. Others have demonstrated higher FA and lower MD in infants and young children^41,42^. Our right-lateralized results are consistent with prior findings showing that prenatal maternal depressive symptoms are associated with prefrontal white matter connections in the right hemisphere^9^. Previous studies have identified right cortex structural variations in children and adolescents with or at risk of depression^43,44^, and EEG studies have demonstrated functional brain activity in infants and children using right frontal electrodes that relate to prenatal anxiety and depression^45^. The right hemisphere is also more implicated in mental health in children^46^ and adolescents^47^, suggesting that functional and structural brain alterations may be an underlying mechanism via which maternal prenatal distress can lead to increased risk of mental health difficulties in offspring.

Importantly, our results also show, for the first time, that social support plays a role not only upon the impact of prenatal distress on pregnant individuals, but also on the developing infant brain. Prenatal distress was negatively related to infant brain functional connectivity in mothers with low social support, but this relationship was not present in individuals with high social support. We found reduced functional connectivity between the right amygdala and superior orbitofrontal cortex was related to higher prenatal distress, suggesting an alteration in the development of the functional connectome that supports emotional regulation and decision making. Previous research in 6-month-old infants demonstrated a positive relationship between amygdala functional connectivity and prenatal depression^48^; negative associations have also been observed in young children^8^. Our data in infants is consistent with the latter finding and demonstrates a similar pattern in infants born to mothers with low social support. Including social support measures and other important covariates may improve the reproducibility of the complex relationship between prenatal distress and the developing infant brain. These findings suggest that social support may disrupt the transmission of prenatal stress to altered functional connectivity in the developing infant brain. This suggests that prenatal social support not only supports the pregnant individual, but also buffers the effects of prenatal maternal distress on infant brain function. Structural connectivity showed alterations associated with prenatal distress, but these were not moderated by social support. It is possible that these relationships may change in later stages of development as the functional and structural connectome of the infant brain continues to develop and refine^40,49,50^.

The current study limitations include self-reported mental health measures and perceived social support. We also relate to indirect imaging measures and further research investigating other regions and networks in the brain as well as emerging behaviours will aid in interpretation. Potential mechanisms linking prenatal maternal distress to infant brain alterations include epigenetic changes such as increased glucocorticoid receptor methylation in children in order to silence stress responses^51^. Invasive animal studies often attribute heightened and sustained cortisol concentrations to changes in infant cognition and behaviour, however the human literature is inconsistent and an indirect mechanism such as regulation of enzymes may be responsible for cortisol metabolism^52^. Prior research has also shown that social support can mitigate the impacts of prenatal stress on this underlying neurophysiology through normalizing the dysregulation of the HPA-axis and consequent cortisol levels^22^, providing further evidence for the importance of social support for pregnant individuals and the developing infant brain.

In conclusion, we show that pregnant individual’s mental health has been especially impacted during the COVID-19 pandemic, and furthermore that heightened prenatal maternal distress can alter the developing infant brain. Our findings suggest long-lasting impacts of this pandemic-related stress on infants, and will help inform health policies and identify families who may benefit from early interventions^53^. The brain shows developmental plasticity in infancy and early childhood, and evidence-based interventions exist to improve children’s behaviours and mental health risk^54^. Importantly, our findings provide evidence that social support can disrupt the intergenerational transmission of stress. This highlights the pressing need for prenatal mental health screening as well as policies that target improving social support^54^, as we have demonstrated that this may support healthy infant brain development in early life.

## Supporting information

eTable1, eTable2

Strobe Checklist

## Acknowledgements

This work was supported by the Alberta Children’s Hospital Research Institute and the Owerko Centre for Neurodevelopment and Mental Health. C.L. receives funding from the Canada Research Chair program. K.Y.M was supported by the T. Chen Fong Postdoctoral Fellowship in Medical Imaging Science. C.L. and K.Y.M. had full access to all the data in the study and takes responsibility for the integrity of the data and the accuracy of the data analysis. K.Y.M. contributed through data collection, analysis, data interpretation, manuscript preparation and revisions. X.L. contributed data collection, analysis, and statistical analyses. D.W. contributed data analysis. L.T.M., G.F.G., and C.L. contributed study conception, funding acquisition, protocol design, and data interpretation. All authors participated in manuscript revisions, approved the final draft, and agree to be accountable for the work. The authors declare no conflicts of interest.

