## Supplementary material for "Prenatal maternal distress during the COVID-19 pandemic and its effects on the infant brain": eTable1, eTable2

**Supplementary Data**

**Supplementary Table 1:** Demographic data

| **Education** | **Percentage of total sample (N = 8602)** |
|---|---|
| Less than high school | 1 |
| High school | 7.7 |
| Trade or Community College | 24.7 |
| University bachelor's degree | 39.5 |
| Master's degree | 19 |
| Doctorate degree | 3.2 |
| Professional (MD, JD, DDS) | 4.8 |
| **Ethnicity** | **Percentage of total sample (N = 8602)** |
| Black | 1.2 |
| Caucasian | 82.7 |
| Chinese | 1.4 |
| First Nations | 1.4 |
| Filipino | 1.2 |
| Hispanic/Latinx | 2.0 |
| Korean | 0.2 |
| Metis | 1.4 |
| South Asian | 2.6 |
| Southeast Asian | 0.4 |
| West Asian | 0.5 |
| Other (not listed above) | 5.1 |

**Supplementary Table 2:** General linear model results relating prenatal maternal distress (PMD) and infant MRI measures (fractional anisotropy (FA), mean diffusivity (MD), and functional connectivity). Significant relationships after FDR correction are shown in bold.

| **MRI Measure** | **Prenatal maternal distress** | | **Maternal Education** | | **Infant Sex** | | **Infant Age** | | **SSEQ** | | **Interaction term** | |
|---|---|---|---|---|---|---|---|---|---|---|---|---|
| | **T** | ***p*** | **T** | ***p*** | **T** | ***p*** | **T** | ***p*** | **T** | ***p*** | **T** | ***p*** |
| Uncinate fasciculus (L) FA | 2.3 | 0.03 | -0.1 | 0.9 | 0.4 | 0.7 | 1.3 | 0.2 | 1.2 | 0.2 | | |
| Uncinate fasciculus (R) FA | **2.8** | **0.008** | 0.4 | 0.7 | 1.7 | 0.1 | 1.6 | 0.1 | 0.6 | 0.5 | | |
| Amygdala-prefrontal (L) FA | 0.8 | 0.4 | -0.5 | 0.6 | 0.7 | 0.5 | 0.9 | 0.4 | 0.3 | 0.8 | | |
| Amygdala-prefrontal (R) FA | 0.8 | 0.4 | 0.9 | 0.9 | 1.8 | 0.08 | 0.7 | 0.5 | -0.1 | 0.9 | | |
| Uncinate fasciculus (L) MD | 2.0 | 0.05 | -0.2 | 0.8 | -0.7 | 0.5 | -1.3 | 0.2 | -0.3 | 0.8 | | |
| Uncinate fasciculus (R) MD | -1.9 | 0.06 | 0.4 | 0.7 | -2.0 | 0.05 | -3.2 | 0.002 | -0.9 | 0.4 | | |
| Amygdala-prefrontal (L) MD | -0.5 | 0.6 | -0.2 | 0.8 | -1.3 | 0.2 | -2.4 | 0.02 | -0.2 | 0.9 | | |
| Amygdala-prefrontal (R) MD | **-2.6** | **0.01** | -0.8 | 0.4 | -1.9 | 0.06 | -1.4 | 0.2 | -0.9 | 0.4 | | |
| Amygdala (L) and superior orbitofrontal cortex (L) connectivity | -1.7 | 0.1 | -1.2 | 0.2 | 0.4 | 0.7 | -3.2 | 0.002 | -1.2 | 0.2 | | |
| Amygdala (R) and superior orbitofrontal cortex (R) connectivity | **-2.7** | **0.01** | -2.9 | 0.006 | 1.8 | 0.08 | -1.6 | 0.1 | -0.7 | 0.5 | **2.3** | **0.02** |
| Amygdala (L) and inferior frontal gyrus (L) connectivity | 0.03 | 0.9 | 0.08 | 0.9 | 0.2 | 0.9 | -1.8 | 0.07 | -0.9 | 0.4 | | |
| Amygdala (R) and inferior frontal gyrus (R) connectivity | 0.4 | 0.7 | -0.9 | 0.3 | 2.1 | 0.04 | -0.9 | 0.4 | -0.4 | 0.7 | | |
